# Evaluation value of bedside ultrasound monitoring of peak flow velocity of abdominal aorta and its branches in volume status of patients with septic shock

**DOI:** 10.1101/2024.02.22.24303100

**Authors:** Chen Wenchong, Guo Weixin, Zeng Xiaodan, Li Deping, Luo Jianli, Zhou Yafeng, Zhao Sicheng, Lu Pinghui

## Abstract

**Objective:** To explore the value of bedside ultrasound in monitoring peak flow velocity of abdominal aorta and its branches in assessing the volume status of patients with septic shock.

**Methods:** A total of 80 patients with septic shock admitted to the Foshan Rehabilitation Hospital(The Fifth People’s Hospital of Foshan) and the Guangdong Provincial People’s Hospital from June 2022 to June 2023 were selected as the research subjects. All patients were treated with mechanical ventilation,and deep venous catheters were placed in the internal jugular vein or subclavian vein to monitor central venous pressure (CVP).PiCCO catheters were placed in the femoral artery to monitor hemodynamic data.At the same time,the maximum internal diameter of the inferior vena cava (IVCmax),the respiratory variation of the inferior vena cava (□IVC),the peak flow velocity of the abdominal aorta (VpeakAA),the peak flow velocity of the celiac artery (VpeakCA),and the peak flow velocity of the superior mesenteric artery (VpeakSMA)were monitored under bedside ultrasound.The global end-diastolic volumn index(GEDI)was used as a grouping indicator,with GEDI≤680ml/m2 as the low-volume group and GEDI≥800ml/m2 as the high-volume group.Compare the differences in peak flow velocity between the abdominal aorta,celiac artery,and superior mesenteric artery between the two groups,and analyze the correlation between peak flow velocity of the abdominal aorta,celiac artery,and superior mesenteric artery and IVCmax,ΔIVC,CVP,and stroke volume variability (SVV);draw a receiver operating characteristic (ROC) curve for the subjects,calculate the area under the curve,and find the diagnostic threshold.

**Results:** There was no significant difference in general data between the two groups (P>0.05).The VpeakAA,VpeakCA,and VpeakSMA in the high-volume group were all higher than those in the low-volume group, and the differences were statistically significant (P<0.05). However,VpeakCA and VpeakSMA were significantly correlated with IVCmax, △ IVC,CVP,and SVV (P<0.05).The ROC curve analysis showed that VpeakAA,VpeakCA,and VpeakSMA could effectively evaluate the volume status of patients with septic shock, and the area under the VpeakSMA curve was 0.846,with a 95% confidence interval of 0.693-0.999,and had high sensitivity and specificity.

**Conclusion:** Bedside ultrasound can dynamically monitor VpeakAA,VpeakCA,and VpeakSMA,which has great value in the evaluation of volume status in patients with septic shock.

---

Hemodynamic instability is one of the important clinical characteristics of patients with septic shock,and circulatory support is the primary diagnostic and treatment strategy for clinical doctors. Research has shown ^[1-2]^ that reasonable fluid resuscitation therapy can effectively protect organ function in critically ill patients and has a positive impact on improving patient prognosis;Unreasonable liquid resuscitation treatment is not only detrimental to the recovery of patients,but can also increase the incidence of complications to a certain extent,which is detrimental to the prognosis of patients. Therefore, accurate evaluation of patient volume responsiveness is particularly crucial before implementing liquid resuscitation treatment for patients ^[3-4]^.Previous studies have found that central venous pressure,inferior vena cava diameter,inferior vena cava respiratory variability,and PiCCO monitoring can all be used to evaluate volume status,but these methods have certain limitations.Considering the redistribution of blood flow within the body and the reduction of gastrointestinal blood flow in a state of insufficient capacity,ensuring blood supply to important organs,monitoring blood flow in the abdominal aorta,abdominal trunk,and superior mesenteric artery may be beneficial for evaluating capacity status.We chose the abdominal aorta and its branches to consider:1.Ultrasound can clearly display;2.There is a possibility of pathophysiology;3.Ultrasound has its fast bedside characteristics.Moreover,arterial ultrasound evaluation has not yet played its role in volume assessment.Therefore,this study aimed to investigate the clinical value of bedside ultrasound monitoring the peak flow velocity of the abdominal aorta,abdominal trunk, and superior mesenteric artery in the assessment of volume status in patients with septic shock.

## 1 Clinical Data and Methods

### 1.1 Clinical Data

A total of 80 patients with septic shock admitted to the Foshan Rehabilitation Hospital(The Fifth People’s Hospital of Foshan) and the Guangdong Provincial People’s Hospital from June 2022 to June 2023 were selected as the research objects,including 48 males and 32 females,aged 45-86 years,with an average of (69.54 ±10.35) years,which met the diagnostic criteria of septic shock^[5]^:(1)There were clear infectious foci;(2)Body temperature is higher than 39 °C or lower than 36°C;(3)Systolic blood pressure is less than 90 mmHg,or the decrease from the original baseline value is more than 40 mmHg,and it lasts for at least 1 hour,or blood pressure can be maintained only by infusion or drugs;(4)Poor tissue perfusion,oliguria for more than 1 hour or acute neurological disorders;(5)It may be found that pathogenic microorganisms grow in blood culture.Exclusion criteria:(1)Previous severe heart,liver, kidney or endocrine system diseases;(2)Patients younger than 18 years old or pregnant;(3)Increased abdominal pressure;(4)Persistent arrhythmia;(5)Shock caused by other factors except infection;(6)Incomplete clinical data.

### 1.2 Methods

All patients were treated with mechanical ventilation.A deep venous catheter was placed in the internal jugular vein or subclavian vein to monitor the central venous pressure(CVP),and a PiCCO catheter was placed in the femoral artery to monitor the hemodynamic data.The global end diastolic volume index (Gedi) was used as the grouping index.Gedi≤680ml/m^2^ was the low volume group,and Gedi≥800ml/m^2^ was the high volume group.The correlation of peak velocity of abdominal aorta, celiac trunk and superior mesenteric artery with the maximum diameter of inferior vena cava, respiratory variability of inferior vena cava,CVP and stroke volume variability (SVV) was analyzed.Draw the receiver operating characteristic(ROC)curve,calculate the area under the curve and find the diagnostic threshold.The study was approved by the Ethics Committee of Foshan Fifth People’s Hospital[2022](No. 0610).The approval date was July 6th, 2022.

### 1.3 Central venous pressure monitoring

A three lumen central venous catheter(American arrow company)was inserted through the right internal jugular vein or the right subclavian vein by Seldinger method.The CVP module (Philips company in the Netherlands) of the pressure sensor and ECG monitor (IntelliVue mp30) was connected.The patient was in a supine position.The height of the pressure sensor was adjusted to be at the same level as the right atrium of the axillary midline.After zero calibration,CVP was read,monitored and recorded.

### 1.4 Pulse indicator continuous cardiac output monitoring cardiac index

PiCCO (pulse indicator continuous coronary output) catheter (pulse company of Germany) was inserted through the right femoral artery by Seldinger method,and connected to the PiCCO module (Philips company of the Netherlands) of the ECG monitor (IntelliVue mp30).The patient’s height,weight and other related information were input,and the body surface area was calculated.Based on the principle of pulmonary thermodilution combined with continuous pulse contour index,15ml of 0□ normal saline was rapidly and evenly injected through the central venous catheter after zero calibration,Then the cardiac index (CI) and other data were obtained,and the average value was obtained after three consecutive measurements.

### 1.5 Bedside ultrasonic monitoring

The bedside ultrasound machine (Philips and Sonosite) was used to monitor the patients.The operation was carried out by the deputy chief physician skilled in ultrasound technology in our hospital.The patients were in the supine position.The vascular ultrasound probe was placed transversely under the xiphoid process,and the abdominal aorta was displayed in the transverse axis direction.Ensure that the image of the abdominal active pulse was placed in the center of the screen,rotate the probe 90° clockwise or counterclockwise,and the abdominal aorta was displayed in the longitudinal axis direction,and then move the probe appropriately.The celiac trunk and superior mesenteric artery were displayed,and then the blood flow spectra of the abdominal aorta,celiac trunk and superior mesenteric artery were displayed according to the ultrasonic standard operation.The peak velocity of the abdominal aorta (VpeakAA), the peak velocity of the celiac trunk artery (VpeakCA) and the peak velocity of the superior mesenteric artery (VpeakSMA) were tested respectively.

### 1.5 Statistical methods

Graphpad prism 8.0 software was used for statistical analysis.The measurement data were expressed as mean ± standard deviation 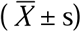 .Paired t-test was used for intra group comparison,and independent sample t-test was used for inter group comparison.The counting data is expressed in (%),and the comparison is made with χ^2^ inspection.Pearson test was used for correlation analysis.Draw the receiver operating characteristic (ROC) curve,calculate the area under the curve and find the diagnostic threshold.The difference was statistically significant (p<0.05).

## 2 Results

### 2.1 All patients were grouped according to Gedi and the comparison of general data between the two groups

All the 80 patients with septic shock, there were 32 patients with Gedi≤680ml/m^2^ (low volume group) and 24 patients with Gedi≥800ml/m^2^ (high volume group).There were no significant differences in age,gender,mean arterial pressure and left ventricular ejection fraction (LVEF) between the two groups (p>0.05).See Table 1.

**Table 1.** Comparison of general data between the two groups.

| Group | Number of cases | Age (years) | Gender (male/female) | Mean arterial pressure (mmHg) | LEVF ( % ) |
| --- | --- | --- | --- | --- | --- |
| low volume group | 32 | $68.37 \pm 13.01$ | 20/12 | $62.32 \pm 10.23$ | $56.43 \pm 8.62$ |
| high volume group | 24 | $70.68 \pm 14.30$ | 14/10 | $64.44 \pm 11.42$ | $57.11 \pm 10.28$ |
| $\chi^2/t$ value | | 0.628 | 0.032 | 0.242 | 0.231 |
| p value |  | 0.576 | 0.765 | 0.864 | 0.776 |

### 2.2 Comparison of peak velocity of abdominal aorta (VpeakAA),peak velocity of abdominal trunk (VpeakCA) and peak velocity of superior mesenteric artery (VpeakSMA) between the two groups

VpeakAA, VpeakCA and VpeakSMA in high-volume group were higher than those in low-volume group,and the differences were statistically significant (p<0.05).See Table 2.

**Table 2.**
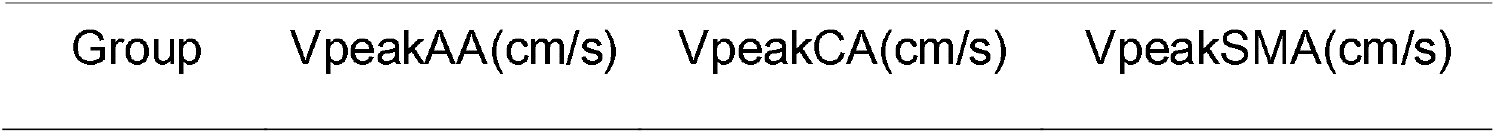
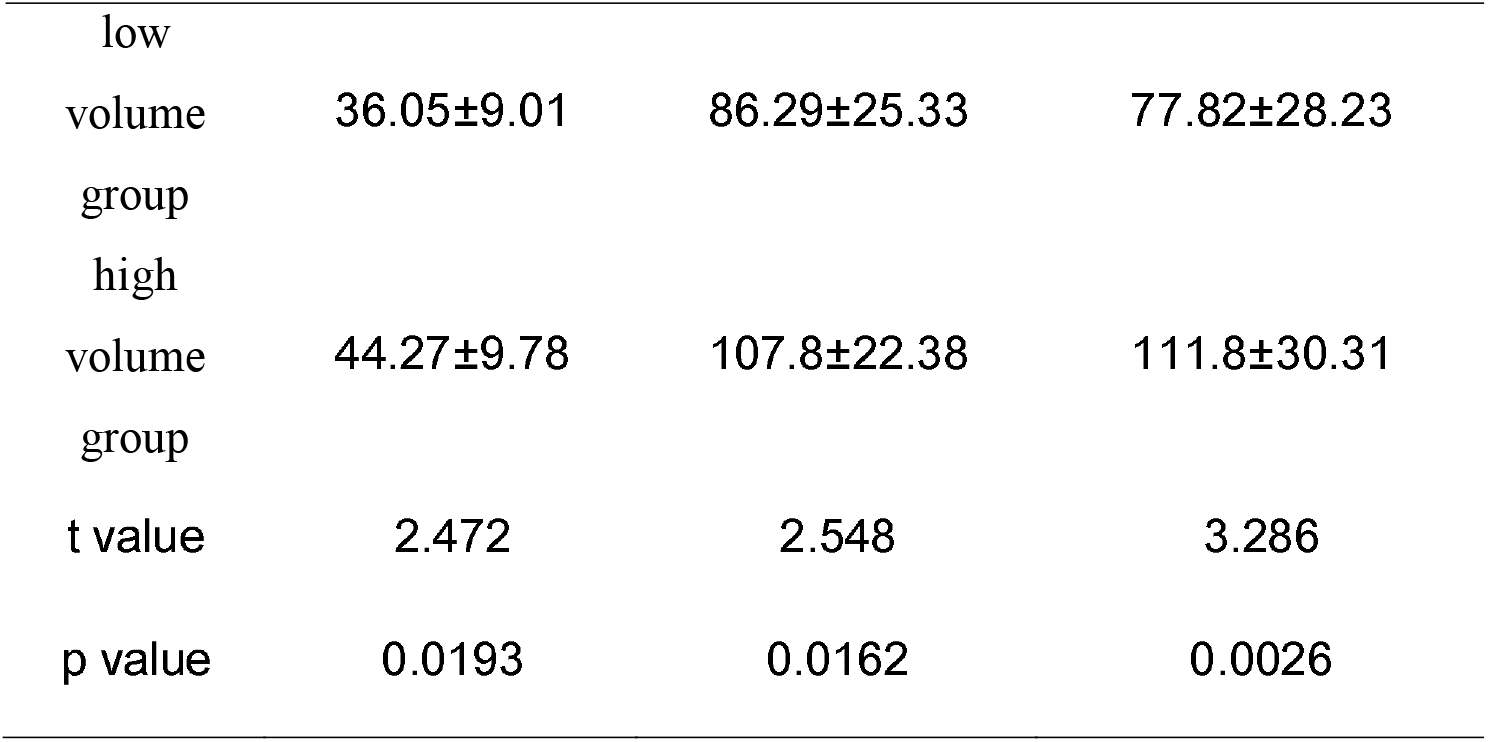
Comparison of VpeakAA, VpeakCA and VpeakSMA between the two groups.

### 2.3 Correlation analysis between VpeakAA and CVP, IVCmax, □ IVC, SVV in two groups

Pearson correlation analysis showed that VpeakAA was positively correlated with CVP (r=0.3173,Fig 1),positively correlated with IVCmax (r=0.2402,Fig 2),negatively correlated with □IVC (r=-0.2920,Fig 3),and negatively correlated with SVV (r=-0.2421,Fig 4),but the differences were not statistically significant (p>0.05).

**Fig 1.**
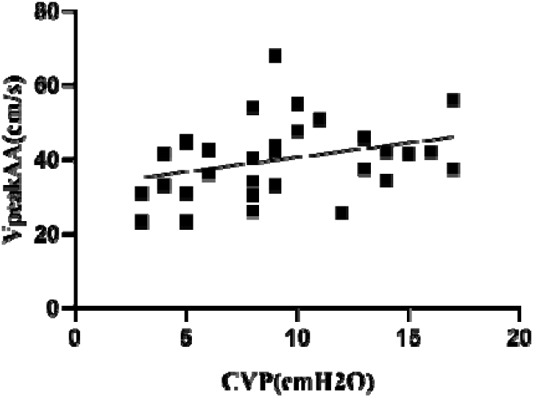
Scatter diagram of VpeakAA and CVP.

**Fig 2.**
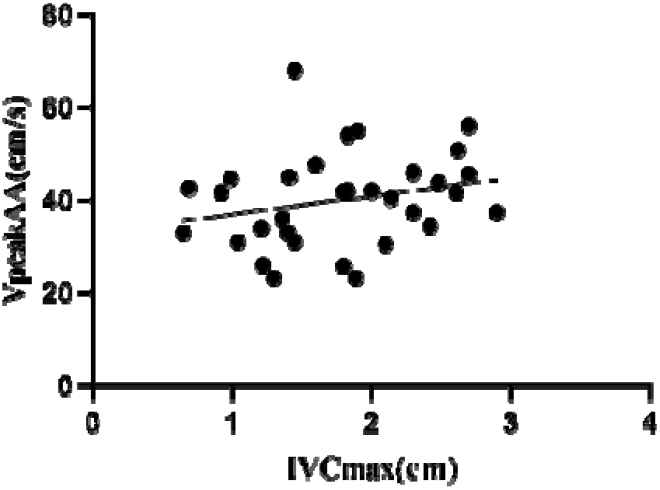
Scatter diagram of VpeakAA and TVCmax.

**Fig 3.**
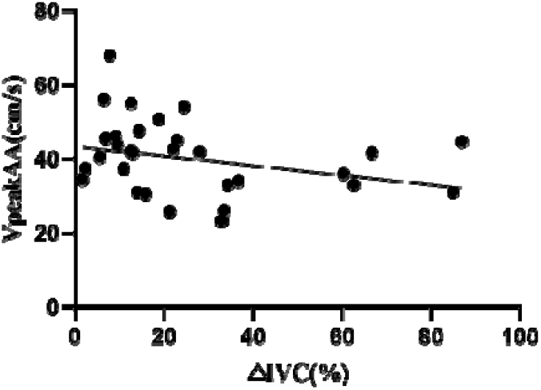
Scatter diagram of VpeakAA and ΔIVC.

**Fig 4.**
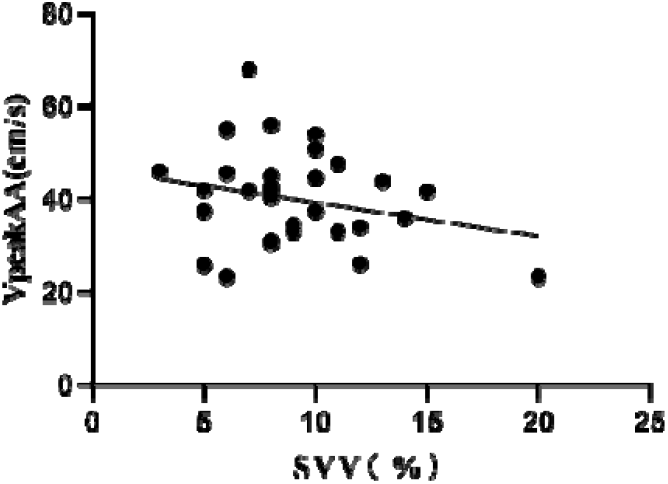
Scatter diagram of VpeakAA and SVV.

### 2.4 Correlation analysis between VpeakCA and CVP,IVCmax,□ IVC,SVV in two groups

Pearson correlation analysis showed that VpeakCA was positively correlated with CVP (r=0.5221,p<0.05) (Fig 5),positively correlated with IVCmax (r=0.3930, p<0.05) (Fig 6),negatively correlated with □ IVC (r=-0.4433,p<0.05) (Fig 7),and negatively correlated with SVV (r=-0.4368,p<0.05) (Fig 8).

**Fig 5.**
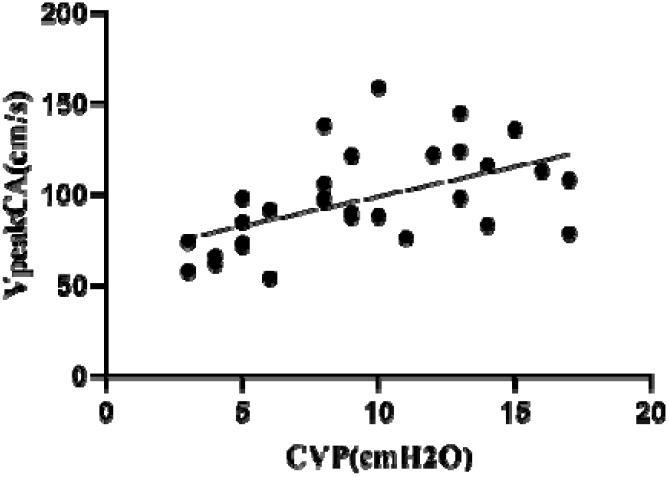
Scatter diagram of VpeakCA and CVP.

**Fig 6.**
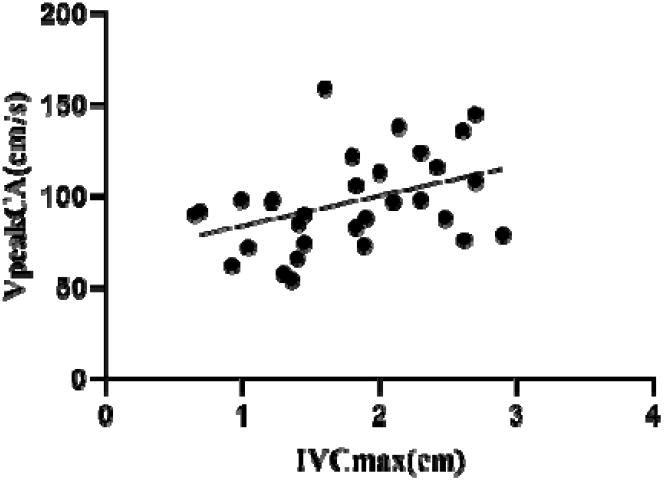
Scatter diagram of VpeakCA and IVCmax.

**Fig 7.**
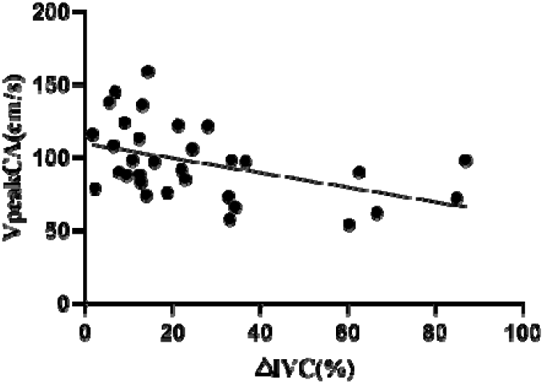
Scatter diagram of VpeakCA and ΔIVC.

**Fig 8.**
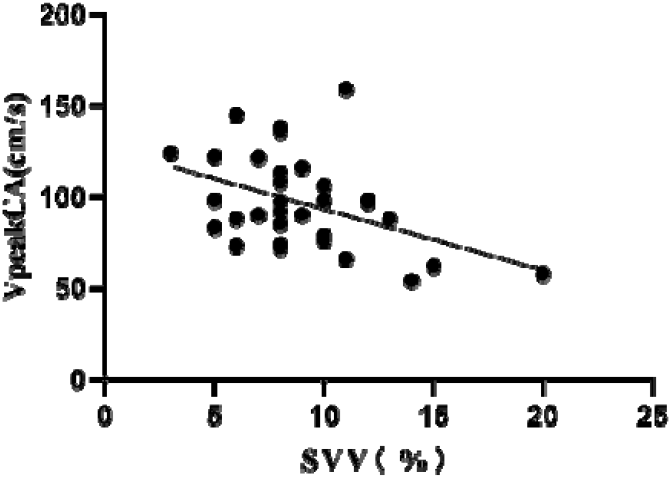
Scatter diagram of VpeakCA and SVV.

### 2.5 Correlation analysis between VpeakSMA and CVP, IVCmax,□IVC,SVV in two groups

Pearson correlation analysis showed that VpeakSMA was positively correlated with CVP (r=0.3665,p<0.05) (Fig 9),positively correlated with IVCmax (r=0.3696, p<0.05) (Fig 10),negatively correlated with □IVC (r=-0.3551,p<0.05) (Fig 11),and negatively correlated with SVV (r=-0.3749,p<0.05) (Fig 12).

**Fig 9.**
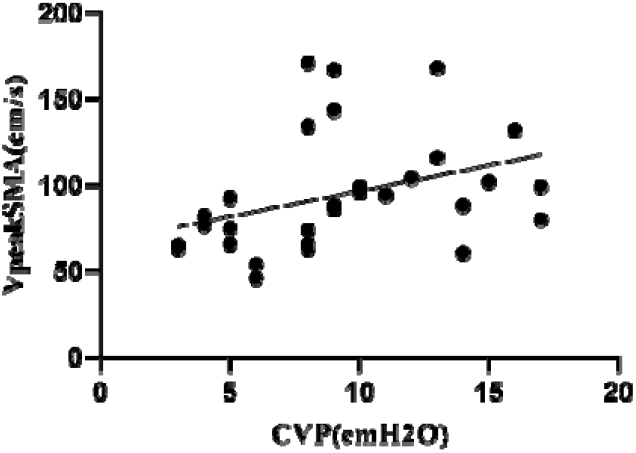
Scatter diagram of VpeakSMA and CVP.

**Fig 10.**
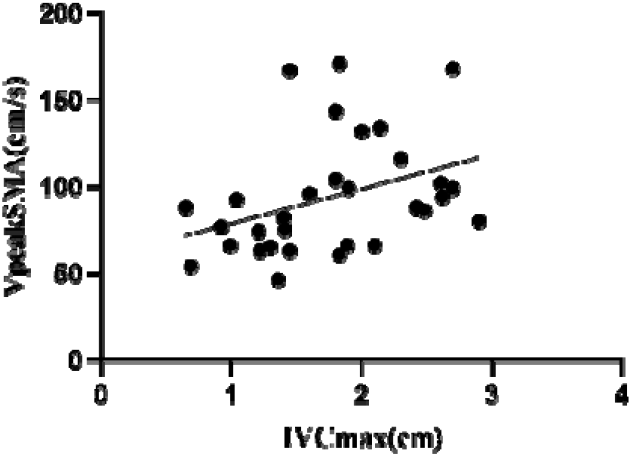
Scatter diagram of VpeakSMA and IVCmax.

**Fig 11.**
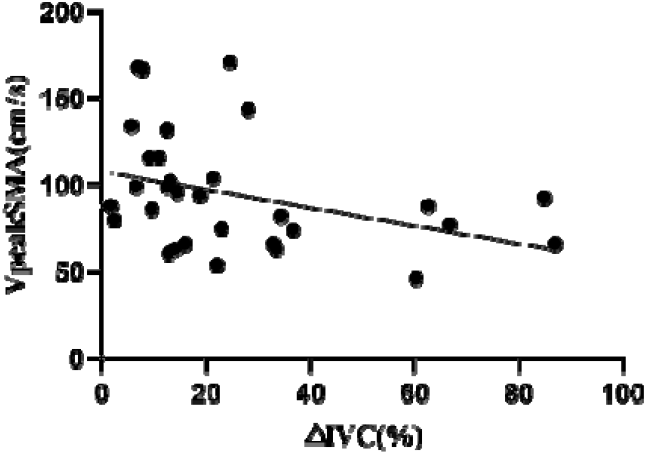
Scatter diagram of VpeakSMA and IVC.

**Fig 12.**
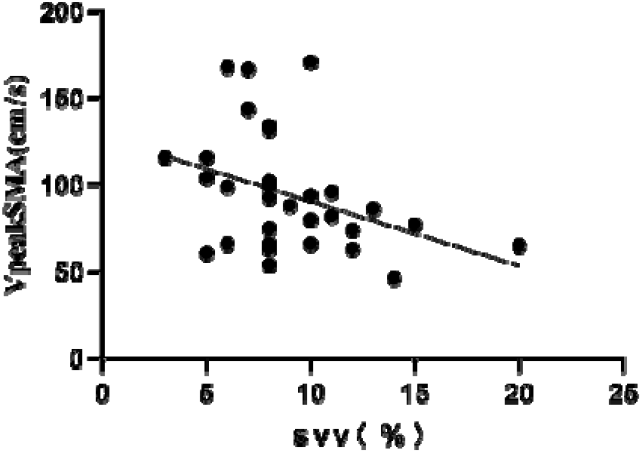
Scatter diagram of VpeakSMA and SVV.

### 2.6 Analysis of receiver operating characteristic (ROC) curve of VpeakAA,VpeakCA and VpeakSMA to assess capacity status

ROC curve analysis showed that VpeakAA,VpeakCA and VpeakSMA could effectively evaluate the volume status of patients with septic shock,and the area under the curve of VpeakSMA was the largest,which was 0.846.Combined with Youden index,when VpeakSMA was 84.1,its sensitivity and specificity were 87.5% and 75.0%,respectively,indicating that VpeakSMA was more valuable in evaluating the volume status of patients with septic shock.See Table 3 and Fig 13.

**Table 3.**
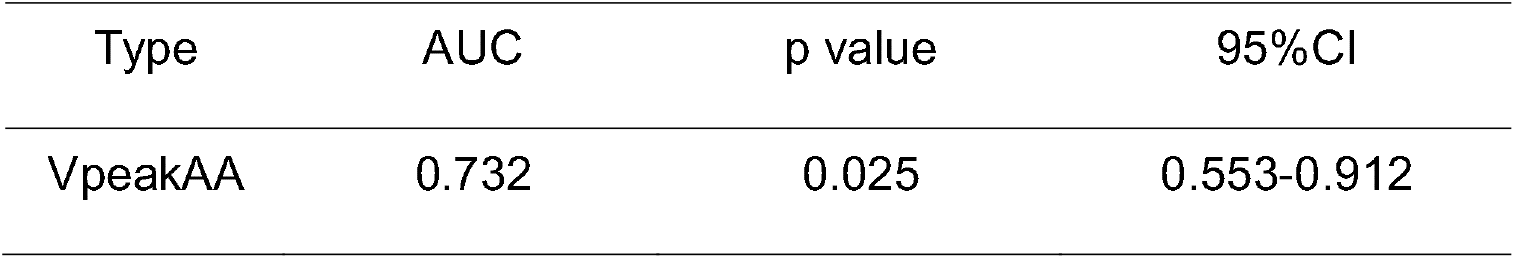
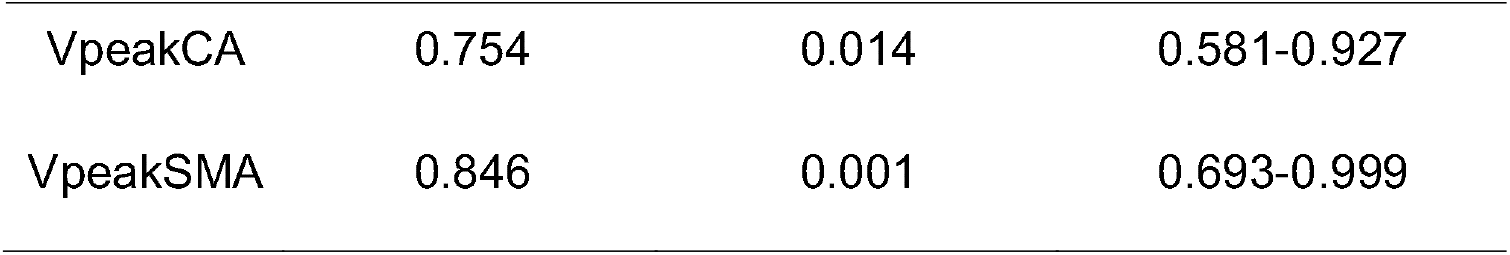
ROC curve analysis of vpeakaa,vpeakca and vpeaksma to assess capacity status.

| Type | AUC | p value | 95%CI |
| --- | --- | --- | --- |
| VpeakAA | 0.732 | 0.025 | 0.553-0.912 |
| VpeakCA | 0.754 | 0.014 | 0.581-0.927 |
| VpeakSMA | 0.846 | 0.001 | 0.693-0.999 |

**Fig 13.**
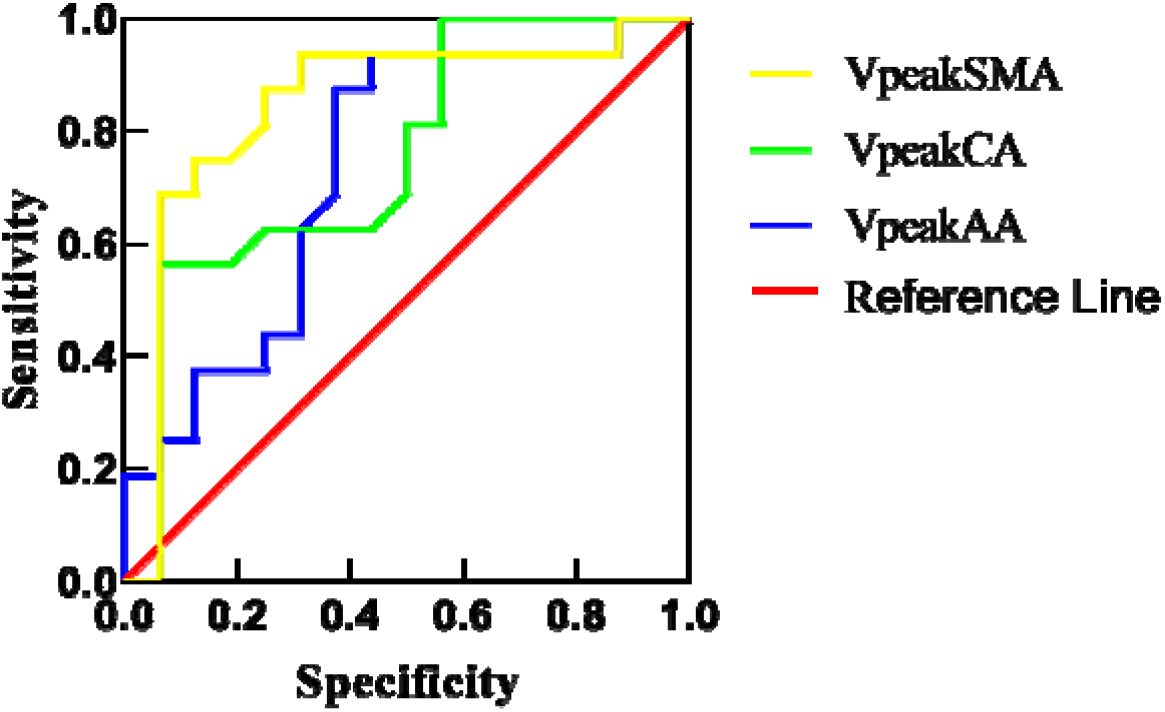
ROC curve of VpeakAA, VpeakCA and VpeakSMA for evaluating capacity status.

## 3 Conclusion

In the past,the capacity load test was often used as the gold standard for evaluating the capacity responsiveness of critically ill patients in clinic.Although the above tests have good accuracy,the test takes a long time,and it is easy to lead to complications such as heart failure and pulmonary edema,with poor safety^[6-7]^.The central venous pressure (CVP) is a classic index to evaluate the state of cardiac preload and guide fluid resuscitation.However, studies have shown that CVP is a pressure index, which can not reflect the preload of the heart.It is not necessarily related to the filling degree of the ventricle,and the central venous pressure is generally too high in patients with right ventricular dysfunction,which is easy to overestimate the fluid volume,resulting in insufficient fluid resuscitation and affecting the prognosis of the disease^[8]^.

In recent years,bedside critical ultrasound has become a hot spot in clinical research.Critical ultrasound is different from traditional diagnostic ultrasound.Critical ultrasound is a dynamic assessment process of problem oriented and multi-objective integration for critical patients by using ultrasound technology under the guidance of critical care medicine theory.It is an important means to determine the direction of critical treatment,especially hemodynamic treatment, and guide fine adjustment^[9]^.Severe ultrasound has the advantages of bedside operability, no radiation,repeatability and low economic cost^[10]^,and it can be checked repeatedly during the treatment process,which is a non-invasive operation.Many studies^[11-13]^ have shown that ultrasound measurement of inferior vena cava diameter,inferior vena cava respiratory variability,left ventricular end diastolic volume,etc.can evaluate the patient’s volume status and volume responsiveness,but the above methods have certain limitations and high requirements for operators,which are easily affected by patient obesity,emphysema,abdominal surgery,tricuspid regurgitation,pulmonary embolism and other factors.The measurement results may have large errors,which will affect the evaluation effect.

Pulse indicated continuous cardiac output (PiCCO) is a minimally invasive,accurate and comprehensive hemodynamic monitoring technology,which plays an accurate guiding role in fluid management.Evaluating the status of circulating volume through measured parameters is a gradually recognized monitoring method in the field of critical care medicine,which can dynamically and effectively evaluate the volume to guide fluid resuscitation^[14]^.However,PiCCO belongs to invasive operation,which requires certain instruments and equipment,and the consumables are expensive.It is not common in non critical departments,and can not be implemented immediately.It has time delay,and there is a certain risk of bloodstream infection,which is not conducive to the extensive clinical development^[15]^.

In the event of septic shock, due to the reduction of effective circulating blood volume and the decrease of blood pressure (mean arterial pressure map<60mmhg),in order to ensure the perfusion of vital organs such as heart and brain,the secretion of catecholamines increases,so as to reduce the perfusion of viscera,skin and other parts first^[16]^.Therefore,in the early stage of blood volume insufficiency,the changes of gastrointestinal blood flow may be earlier than the changes of inferior vena cava and left ventricular end diastolic volume.Monitoring the changes of gastrointestinal blood flow may help to evaluate the volume status of critically ill patients earlier.In this study,the VpeakAA, VpeakCA and VpeakSMA in patients with septic shock were measured by bedside ultrasound, and the CVP, IVCmax, □IVC and PiCCO were compared.The results showed that VpeakAA, VpeakCA and VpeakSMA were significantly correlated with the above indicators of traditional assessment of volume status.The ROC curve analysis showed that VpeakAA, VpeakCA and VpeakSMA could effectively evaluate the volume status of patients with septic shock,and the area under the curve of VpeakSMA was the largest,suggesting that VpeakSMA was more valuable in volume evaluation.

In conclusion,bedside ultrasound can dynamically monitor the peak flow velocity of abdominal aorta,celiac trunk and superior mesenteric artery due to its advantages of noninvasive,simple and rapid operation and good repeatability.It has great value in the volume assessment of patients with septic shock, providing an alternative or even better choice for volume assessment,guiding fluid resuscitation more accurately and improving the prognosis of patients.However,the sample size of this study is small,and there may be human errors in the ultrasonic monitoring,which may lead to the deviation of the results.In the next step,we need to further increase the sample size for more in-depth research,and further standardize the ultrasonic monitoring methods,so as to achieve more accurate research results.

## Data Availability

All data produced in the present work are contained in the manuscript

